# The management of blunt pancreatic injury in children in New South Wales

**DOI:** 10.1101/2023.07.26.23292584

**Authors:** Sally T W Chung, Andrew J A Holland, Julie Brown, Susan E Adams

## Abstract

**Background:** Blunt pancreatic injury is uncommon but associated with considerable morbidity. The optimal management strategy for children with this injury remains unclear, with laparotomy rates in North America of up to 55%. This has not been studied at a population level in Australia. This study aimed to examine the management of children with blunt pancreatic injury in New South Wales (NSW), Australia.

**Methods:** Using the NSW Admitted Patient Data Collection from 2001 to 2019, we identified patients <16 years old with blunt pancreatic injury. Cases were categorised as undergoing non-operative or operative management. Patient and injury characteristics and outcomes were compared between non-operative and operative groups. Independent predictors of operative management were identified using multivariable logistic regression.

**Results:** 139 cases were identified: 37 (26.6%) underwent operative management, of whom 11 (29.7%) had a pancreas-specific operation. Two-thirds were managed at a paediatric trauma centre. Operation rates were highest in adult trauma centres, although treatment outside a paediatric trauma centre overall was not associated with operative management. Independent predictors of operative management were high-grade pancreatic injury, hollow viscus injury and transfusion. Morbidity and mortality were 30.2% and 1.4%, respectively.

**Conclusion:** Blunt pancreatic injury continues to carry substantial morbidity. Operation rates in NSW are lower than those reported in North America, with similar outcomes. Unlike other solid organ injuries, most children are managed at paediatric trauma centres. Future studies should investigate factors driving management decisions in the Australian context, with the aim of developing paediatric guidelines promoting non-operative management.

**Highlights:** Operation rates for children with BPI were much lower than reported in international studies, with comparable morbidity and lower mortality.

Operation rates were highest at ATCs, although, overall, children with BPI managed outside PTCs were not more likely to undergo an operation.

Penetrating pancreatic injury is rare compared to the US, likely reflecting that firearm-related injury in Australia remains uncommon.

## 1. Introduction

Worldwide, traumatic injury is the leading cause of mortality in children over the age of one and in adults up to the age of 45 [1–3]. Pancreatic injury in children is less common than injury to other solid organs [4], occurring in 0.6% of blunt abdominal trauma [5]. However, it is associated with significant morbidity, including pseudocyst, fistula formation, pancreatitis and sepsis [5–12]. Furthermore, there remains a mortality rate in children of 3.1-5.3% [5, 7]. Pancreatic injury can be caused by blunt or penetrating trauma, with blunt pancreatic injury (BPI) predominating amongst children [5].

In 1989, the American Association for the Surgery of Trauma (AAST) published the Organ Injury Scale (OIS) to describe traumatic injury severity [13]. Pancreatic injury grade is determined by the integrity of the main pancreatic duct and the location of injury. Grade I and II injuries have no ductal involvement and are considered low-grade. Grades III-V involve the pancreatic duct and are considered high-grade, being associated with increased morbidity and mortality [14]. Grading of pancreatic injury is useful in guiding management and predicting patient outcomes [14].

In haemodynamically stable patients, non-operative management (NOM) of other abdominal solid organ injuries has become the standard of care for both children and adults [15–18]. In contrast, the optimal management strategy for pancreatic injury remains unclear, with ongoing controversy over the benefits of NOM versus operative management (OM). There are guidelines for the management of adults with pancreatic injury. NOM is supported for low-grade pancreatic injury if there are no other indications for exploratory laparotomy, due to low morbidity [19, 20]. For grade III and IV injuries – which involve distal or proximal duct disruption, respectively, OM is recommended [19, 20]. There is no consensus for the management of grade V injury, which involves massive disruption of the pancreatic head [19]. However, the evidence for these guidelines comes from observational, often retrospective studies of both blunt and penetrating pancreatic injury, and many recommendations are conditional given the low power, heterogeneity and limited reported outcomes [19].

Conversely, there are no clear management guidelines for children with BPI. A 2014 Cochrane review concluded that whilst consensus for NOM of low-grade BPI exists, management of high-grade BPI remains controversial [21, 22]. A more recent systematic review confirmed that NOM is justified for low-grade BPI, with no significant differences in mortality or pseudocyst formation compared to OM [22]. For children with high-grade BPI, there were also no mortality differences between management groups, although significantly more pseudocysts occurred in those managed non-operatively [22]; even so, pseudocysts can often be managed non-operatively [23, 24]. NOM is generally now preferred even for proximal high-grade injuries in children given the significant morbidity associated with pancreaticoduodenectomy [25]. Studies have shown a trend towards NOM for all BPI and specifically for high-grade injury without any changes in outcomes [26, 27]. Compared to children, literature reporting successful NOM of high-grade pancreatic injury amongst adults is sparse [28–31]. Whilst there is an overall trend towards NOM [32], rates remain considerably lower than in children.

Trauma systems are designed to coordinate and optimise a multidisciplinary response to the injured patient, encapsulating pre-hospital care, acute hospital care and post-injury rehabilitation. It is widely recognised that they reduce mortality and improve outcomes [33–36]. New South Wales, a state within Australia with a population of 8.17 million people, has a trauma system consisting of three PTCs, eight metropolitan ATCs and 10 regional trauma centres (RTCs). However, there are no set trauma triage criteria for children, and many injured children are managed in non-paediatric settings. For example, only one-third of children with blunt splenic injury in NSW are admitted to or transferred to a PTC [37]. This is important because there is evidence that children who are managed at a paediatric trauma centre (PTC) rather than an adult trauma centre (ATC) have lower mortality rates, shorter length of stays and improved functional outcomes [38–43]. The blunt splenic injury intervention rate is a key quality improvement metric for paediatric trauma [44]: studies in New South Wales (NSW) [37] and elsewhere [45–48], have shown significantly lower splenectomy rates when children with blunt splenic injury are managed in PTCs [13, 14], indicating improved paediatric trauma care.

To our knowledge, BPI has not been studied at a population level for children in Australia. There is a need for current knowledge on the epidemiology, management and outcomes of children with BPI in NSW. The results from this study will help inform injury prevention strategies, health professional education and trauma service planning.

## 2. Study aims

In children aged 0-15 years with BPI, our study aimed to 1) describe patient demographics and injury characteristics, 2) analyse management strategies and determine independent predictors of OM and 3) describe short-term outcomes.

## 3. Methodology

This study was approved by the NSW Population and Health Services Research Ethics Committee (Approval number: 2019/ETH00387/48707). This manuscript was written following the recommendations of the Strengthening the Reporting of Observational Studies in Epidemiology (STROBE) Statement [49].

### 3.1 Study design

This was a retrospective cohort data linkage study including children aged 0-15 who were admitted to an NSW hospital with BPI.

### 3.2 Study population

#### 3.2.1 Hospital and Mortality data

The Admitted Patient Data Collection (APDC) was the primary data source. This database is maintained by the NSW Ministry of Health and contains information on all NSW hospital admissions, including data on demographics, facilities, diagnoses, external causes of injury, procedures and discharge destinations. Diagnoses, external causes and procedures are coded according to the International Classification of Diseases, Tenth revision, Australian Modification (ICD-10-AM). Mortality data were obtained from the Registry of Births, Deaths and Marriages and the Australian Bureau of Statistics deaths registration and linked to APDC data by the NSW Centre for Health Record Linkage. All data were provided in a de-identified format.

Each health record in the APDC reflects an individual episode of care in a hospital, which ends with the discharge, transfer or death of the patient, or when the patient’s service category changes. Thus, a single injury may engender multiple episodes of care if the patient was transferred to a different hospital (e.g., from a metropolitan local hospital to a PTC). All episodes of care with ≤1-day difference between discharge and subsequent admission were treated as being related to the original pancreatic injury and linked to form a single ‘period of care’ (i.e., all episodes of care related to the pancreatic injury until discharge from the health system). Where there was a break from the health system of >1 day, subsequent records were treated as readmissions.

#### 3.2.2 Final study population

The dataset was evaluated between 2001-2019 for patients aged 0-15 with a diagnosis of pancreatic injury. Those with penetrating pancreatic injury were subsequently excluded from analysis. Data extraction and creation of explanatory and outcome variables were conducted in R version 4.05 (R Foundation for Statistical Computing, Vienna, Austria).

### 3.3 Explanatory variables

Variables were created for age, gender, socioeconomic status, hospital type, intent of injury, mechanism of injury, location of pancreatic injury, injury severity, associated injuries and the need for transfusion. Appendix A details explanatory variables and their definitions.

Pancreatic injury grade was classified as ‘high-grade’ or ‘low-grade’: injury to ‘multiple or other parts’ of the pancreas (*ICD-10-AM: S36.29)* was used as a proxy for high-grade injury, with this category inclusive of pancreatic duct injury, whereas injury to individual parts (head, body, tail) *(ICD-10-AM: S 36.21, S36.22, S36.23)* or ‘unspecified’ parts of the pancreas *(ICD-10-AM: S36.20)* was classified as low-grade injury. Overall injury severity was represented with the International Classification of Disease (ICD)-based Injury Severity Score (ICISS), calculated using paediatric Survival Risk Ratios (SRRs) derived by Mitchell et al [50]. An SRR is defined as the ratio of all survivors with a particular ICD-10-AM injury code to all patients with that injury code, including deaths. The ICISS is calculated by multiplying all the paediatric SRRs for each of the patient’s injuries. It ranges from 0-1, with a lower number indicating greater injury severity.

### 3.4 Outcomes

The primary outcome was management strategy: operative management (OM: any laparotomy or laparoscopy) and non-operative management (NOM: no laparotomy or laparoscopy). In those undergoing OM, we further examined whether a pancreas-specific operation (primary pancreatic resection or repair) occurred. Notably, our definition of OM included those operations without a pancreas-specific operation. Secondary outcomes were pancreatic complications, other complications, hospital length of stay (LOS), intensive care unit (ICU) LOS, hours on mechanical ventilation, mortality within 90 days and readmission within 90 days. Outcomes are summarised in Appendix B.

### 3.5 Statistical analysis

Categorical variables were reported using frequency and percentages and continuous variables using median and interquartile ranges (IQRs). Categorical variables were compared using the Chi-squared or Fisher’s exact test and continuous variables using the Mann-Whitney-U test. Multivariable logistic regression was used to identify independent predictors of OM after adjusting for other demographic and injury characteristics in children. Explanatory variables for inclusion in the initial model were chosen *a priori* based on clinical relevance and published literature. Those with insufficient numbers were coalesced with other related variables or excluded. We used a backwards stepwise method with likelihood ratios, retaining variables if they had a p-value of <0.1 or were deemed clinically relevant. Variables believed to be the strongest predictors of OM or of particular interest were forced into the model regardless of p-value. Multicollinearity was assessed using the variance inflation factor, and the Akaike Information Criteria (AIC) was used to assess model performance. Odds ratios (OR) and 95% Confidence Intervals (CI) were calculated. A p-value of <0.05 was considered statistically significant. SPSS Statistics Version 26 (IBM, Armonk, NY, USA) was used for all analyses.

## 4. Results

Between 2001-2019, 145 children were admitted to an NSW hospital with a pancreatic injury. Penetrating pancreatic injury was present in six (4.1%) children; firearms were responsible for one of these cases. After excluding these cases, 139 patients with BPI were available for analysis.

### 4.1 Patient demographics and injury characteristics

Table 1 summarises the patient demographics and injury characteristics, stratified by NOM (n=102/139, 73.4%) versus OM (n=37/139, 26.6%). Overall, children with BPI had a median age of 11.1 years: around two-thirds were male. Two-thirds were managed at a PTC (n=91/139, 65.5%), just over 20% at a non-paediatric trauma centre (ATC: n=12, 8.6%; RTC: n=17, 12.2%) and the remainder (n= 19, 13.7%) away from any trauma centre. Most injuries were accidental, with pedal cyclist (n=36/139, 25.9%) and motor vehicle accident (MVA, n=24/139, 17.3%) as the most common injury mechanisms. Almost a third of injuries were high-grade (n=41/139, 29.5%) and the pancreatic tail was the most common single site of injury (n=18/139, 12.9%). Just over two-thirds (n=85/139, 68.3%) of children had associated injuries and 60.4% (n=84/139) had other intraabdominal injuries.

**Table 1.**
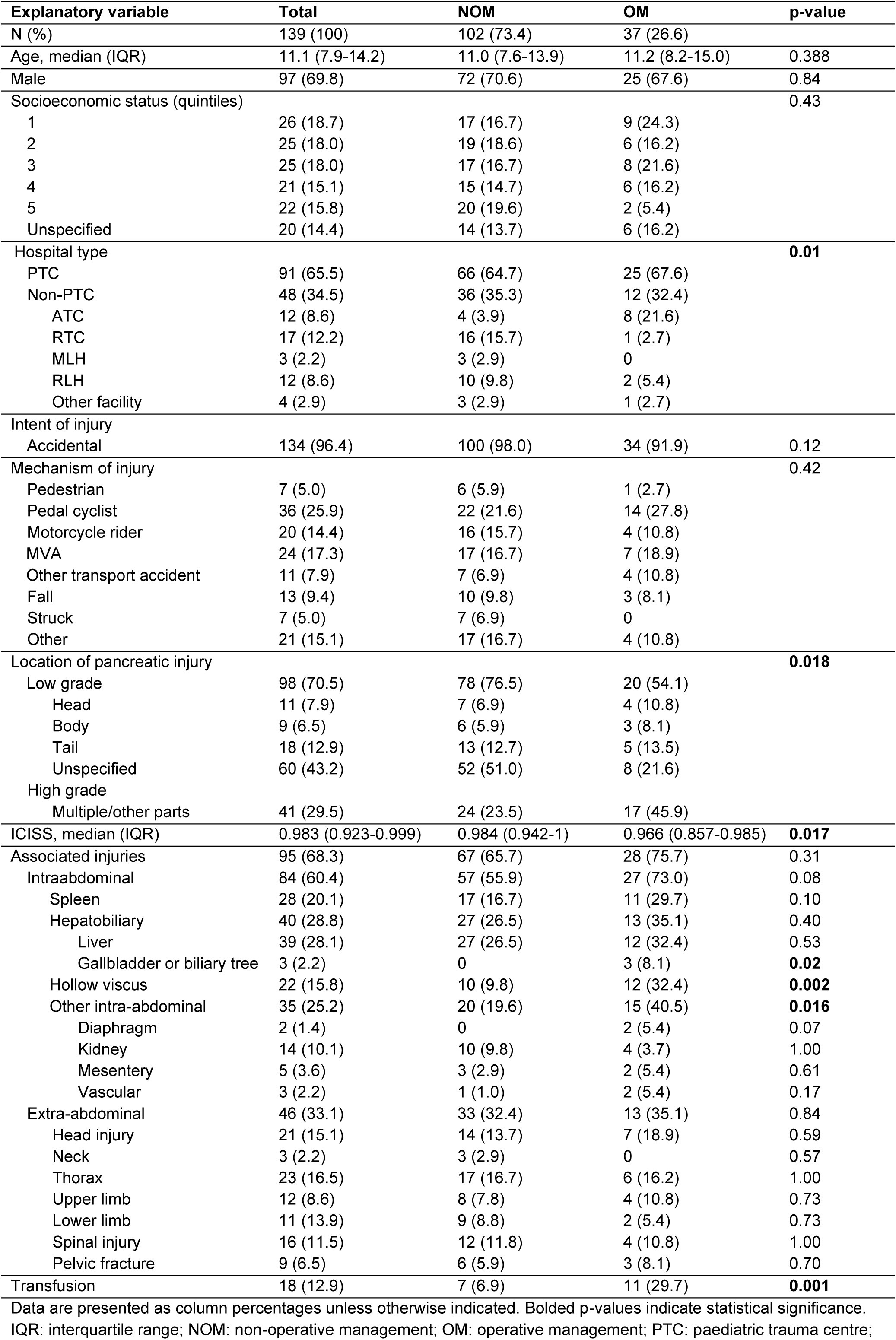
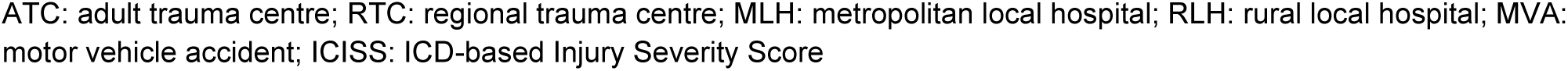
Patient demographics and injury characteristics by management strategy.

| Explanatory variable | Total | NOM | OM | p-value |
| --- | --- | --- | --- | --- |
| N (%) | 139 (100) | 102 (73.4) | 37 (26.6) |  |
| Age, median (IQR) | 11.1 (7.9-14.2) | 11.0 (7.6-13.9) | 11.2 (8.2-15.0) | 0.388 |
| Male | 97 (69.8) | 72 (70.6) | 25 (67.6) | 0.84 |
| Socioeconomic status (quintiles) |  |  |  | 0.43 |
| 1 | 26 (18.7) | 17 (16.7) | 9 (24.3) |  |
| 2 | 25 (18.0) | 19 (18.6) | 6 (16.2) |  |
| 3 | 25 (18.0) | 17 (16.7) | 8 (21.6) |  |
| 4 | 21 (15.1) | 15 (14.7) | 6 (16.2) |  |
| 5 | 22 (15.8) | 20 (19.6) | 2 (5.4) |  |
| Unspecified | 20 (14.4) | 14 (13.7) | 6 (16.2) |  |
| Hospital type |  |  |  | **0.01** |
| PTC | 91 (65.5) | 66 (64.7) | 25 (67.6) |  |
| Non-PTC | 48 (34.5) | 36 (35.3) | 12 (32.4) |  |
| ATC | 12 (8.6) | 4 (3.9) | 8 (21.6) |  |
| RTC | 17 (12.2) | 16 (15.7) | 1 (2.7) |  |
| MLH | 3 (2.2) | 3 (2.9) | 0 |  |
| RLH | 12 (8.6) | 10 (9.8) | 2 (5.4) |  |
| Other facility | 4 (2.9) | 3 (2.9) | 1 (2.7) |  |
| Intent of injury |  |  |  |  |
| Accidental | 134 (96.4) | 100 (98.0) | 34 (91.9) | 0.12 |
| Mechanism of injury |  |  |  | 0.42 |
| Pedestrian | 7 (5.0) | 6 (5.9) | 1 (2.7) |  |
| Pedal cyclist | 36 (25.9) | 22 (21.6) | 14 (27.8) |  |
| Motorcycle rider | 20 (14.4) | 16 (15.7) | 4 (10.8) |  |
| MVA | 24 (17.3) | 17 (16.7) | 7 (18.9) |  |
| Other transport accident | 11 (7.9) | 7 (6.9) | 4 (10.8) |  |
| Fall | 13 (9.4) | 10 (9.8) | 3 (8.1) |  |
| Struck | 7 (5.0) | 7 (6.9) | 0 |  |
| Other | 21 (15.1) | 17 (16.7) | 4 (10.8) |  |
| Location of pancreatic injury |  |  |  | **0.018** |
| Low grade | 98 (70.5) | 78 (76.5) | 20 (54.1) |  |
| Head | 11 (7.9) | 7 (6.9) | 4 (10.8) |  |
| Body | 9 (6.5) | 6 (5.9) | 3 (8.1) |  |
| Tail | 18 (12.9) | 13 (12.7) | 5 (13.5) |  |
| Unspecified | 60 (43.2) | 52 (51.0) | 8 (21.6) |  |
| High grade |  |  |  |  |
| Multiple/other parts | 41 (29.5) | 24 (23.5) | 17 (45.9) |  |
| ICISS, median (IQR) | 0.983 (0.923-0.999) | 0.984 (0.942-1) | 0.966 (0.857-0.985) | **0.017** |
| Associated injuries | 95 (68.3) | 67 (65.7) | 28 (75.7) | 0.31 |
| Intraabdominal | 84 (60.4) | 57 (55.9) | 27 (73.0) | 0.08 |
| Spleen | 28 (20.1) | 17 (16.7) | 11 (29.7) | 0.10 |
| Hepatobiliary | 40 (28.8) | 27 (26.5) | 13 (35.1) | 0.40 |
| Liver | 39 (28.1) | 27 (26.5) | 12 (32.4) | 0.53 |
| Gallbladder or biliary tree | 3 (2.2) | 0 | 3 (8.1) | **0.02** |
| Hollow viscus | 22 (15.8) | 10 (9.8) | 12 (32.4) | **0.002** |
| Other intra-abdominal | 35 (25.2) | 20 (19.6) | 15 (40.5) | **0.016** |
| Diaphragm | 2 (1.4) | 0 | 2 (5.4) | 0.07 |
| Kidney | 14 (10.1) | 10 (9.8) | 4 (3.7) | 1.00 |
| Mesentery | 5 (3.6) | 3 (2.9) | 2 (5.4) | 0.61 |
| Vascular | 3 (2.2) | 1 (1.0) | 2 (5.4) | 0.17 |
| Extra-abdominal | 46 (33.1) | 33 (32.4) | 13 (35.1) | 0.84 |
| Head injury | 21 (15.1) | 14 (13.7) | 7 (18.9) | 0.59 |
| Neck | 3 (2.2) | 3 (2.9) | 0 | 0.57 |
| Thorax | 23 (16.5) | 17 (16.7) | 6 (16.2) | 1.00 |
| Upper limb | 12 (8.6) | 8 (7.8) | 4 (10.8) | 0.73 |
| Lower limb | 11 (13.9) | 9 (8.8) | 2 (5.4) | 0.73 |
| Spinal injury | 16 (11.5) | 12 (11.8) | 4 (10.8) | 1.00 |
| Pelvic fracture | 9 (6.5) | 6 (5.9) | 3 (8.1) | 0.70 |
| Transfusion | 18 (12.9) | 7 (6.9) | 11 (29.7) | **0.001** |

Data are presented as column percentages unless otherwise indicated. Bolded p-values indicate statistical significance.
IQR: interquartile range; NOM: non-operative management; OM: operative management; PTC: paediatric trauma centre;
ATC: adult trauma centre; RTC: regional trauma centre; MLH: metropolitan local hospital; RLH: rural local hospital; MVA: motor vehicle accident; ICISS: ICD-based Injury Severity Score

The OM and NOM groups were similar in age, sex, socioeconomic status and mechanisms of injury. However, children who had OM were more likely to have a gall bladder or biliary tree injury (p=0.02), hollow viscus injury (p=0.002) and a transfusion (p=0.001). 73% (n=27/37) of children undergoing OM had associated intraabdominal injuries. On further analysis, OM occurred in one-fifth of low-grade injury compared to two-fifths of high-grade injury (n=20/98, 20.4% vs n=17/41, 41.5%; p=0.01).

### 4.2 Operative or interventional procedures

Table 2 summarises specific operative and interventional procedures. Just over a quarter (n=37/139, 26.6%) underwent OM and 7.9% (n=11/139) had a pancreas-specific operation. The overall splenectomy rate was 2.2%. Of the OM group, 29.7% (n=11/37) had a pancreas-specific operation. Furthermore, 29.7% (n=11/37) of those undergoing OM had a concomitant splenic injury, however, splenectomy was performed in only three. Children who underwent OM were also more likely to need total parenteral nutrition (p=0.001).

**Table 2.** Operative or interventional procedures by management strategy in children.

| **Procedure** | **Overall** | **NOM** | **OM** | **p-value** |
| --- | --- | --- | --- | --- |
| N (%) | 139 (100) | 102 (73.4) | 37 (26.6) |  |
| Exploratory laparotomy | 5 (3.6) | 0 | 5 (13.5) | - |
| Diagnostic laparoscopy | 2 (1.4) | 0 | 2 (5.4) | - |
| **Pancreatic operation:** | 11 (7.9) | 0 | 11 (29.7) | - |
| Pancreatic resection | 9 (6.5) | 0 | 9 (24.3) | - |
| Pancreatic repair | 2 (1.4) | 0 | 2 (5.7) | - |
| Pancreatic drainage | 8 (5.8) | 0 | 8 (21.6) | - |
| Pancreatic exploration | 0 (0) | 0 | 0 (0) | - |
| Diaphragm repair | 1 (0.7) | 0 | 1 (2.7) | - |
| Splenectomy | 3 (2.2) | 0 | 3 (8.1) | - |
| Spleen-preserving procedure | 0 | 0 | 0 (0) | - |
| Liver operation | 1 (0.7) | 0 | 1 (2.7) | - |
| Gallbladder or biliary tract operation | 10 (7.2) | 0 | 10 (27.0) | - |
| Gastric operation | 2 (1.4) | 0 | 2 (5.4) | - |
| Small bowel operation | 4 (2.9) | 0 | 4 (10.8) | - |
| Colonic operation | 3 (2.2) | 0 | 3 (8.1) | - |
| Bladder operation | 0 | 0 | 0 | - |
| Kidney operation | 2 (1.4) | 0 | 2 (5.4) | - |
| Other abdominal operation | 17 (12.2) | 0 | 17 (45.9) | - |
| Endoscopic procedure | 4 (2.9) | 1 (1.0) | 3 (8.1) | 0.58 |
| TPN | 16 (11.5) | 6 (5.9) | 10 (27.0) | **0.001** |

Data are presented as column percentages unless otherwise indicated. Bolded p-values indicate statistical significance. NOM: non-operative management; OM: operative management; TPN: total parenteral nutrition

Over 90% (n=34/37, 91.9%) of children undergoing OM were managed at any trauma centre. Specifically, OM was most common at ATCs (n=8/12, 66.7%). This compares to OM in 26/108 (24.1%) of cases at non-paediatric trauma centres (p=0.004). By age group, OM was commonest in children under the age of four (n=5/15, 33.3%), all of whom were operated at PTCs, followed by those aged 13-15 years (n=16/54, 29.6%). In these older children, most operations were undertaken at PTCs (n=8/27, 29.6%) and ATCs (n=5/9, 55.6%) (Table 3).

**Table 3.** The relationship between age group, hospital type and management strategy in children with blunt pancreatic injury.

**a. Management strategy according to age subgroup**
| Age subgroup | Total | NOM | OM |
| --- | --- | --- | --- |
| N | 139 | 102 (68.2) | 37 (31.8) |
| <4 | 15 (5.4) | 10 (66.7) | 5 (33.3) |
| 4-6 | 11 (3.9) | 9 (81.8) | 2 (18.2) |
| 7-9 | 28 (10.0) | 23 (81.8) | 5 (18.2) |
| 10-12 | 31 (11.1) | 22 (71.0) | 9 (29.0) |
| 13-15 | 54 (19.3) | 38 (70.4) | 16 (29.6) |

**b. Management strategy by age subgroup and hospital type**
| Age subgroup | PTC |  | ATC |  | RTC |  | MLH |  | RLH |  | Other facility |  |
| --- | --- | --- | --- | --- | --- | --- | --- | --- | --- | --- | --- | --- |
|  | NOM | OM | NOM | OM | NOM | OM | NOM | OM | NOM | OM | NOM | OM |
| <4 | 9 (64.3) | 5 (35.7) | 0 | 0 | 1 (100) | 0 | 0 | 0 | 0 | 0 | 0 | 0 |
| 4-6 | 5 (71.4) | 2 (28.6) | 0 | 0 | 3 (100) | 0 | 0 | 0 | 1 (100) | 0 | 0 | 0 |
| 7-9 | 18 (85.7) | 3 (14.3) | 0 | 1 (100) | 0 | 0 | 1 (100) | 0 | 3 (75.0) | 1 (25.0) | 1 (100) | 0 |
| 10-12 | 15 (68.2) | 7 (31.8) | 0 | 2 (100) | 6 (100) | 0 | 0 | 0 | 1 (100) | 0 | 0 | 0 |
| 13-15 | 19 (70.4) | 8 (29.6) | 4 (44.4) | 5 (55.6) | 6 (85.7) | 1 (14.3) | 2 (100) | 0 | 5 (83.3) | 1 (16.7) | 2 (66.7) | 1 (33.3) |

Age subgroups are in years. Data are presented as percentages of all patients in that age subgroup managed at that hospital type (see Appendix C for table of total number of patients in each age subgroup managed at each hospital). NOM: non-operative management; OM: operative management; PTC: paediatric trauma centre; ATC: adult trauma centre; RTC: regional trauma centre; MLH: metropolitan local hospital; RLH: rural local hospital. Percentages represent the per cent of patients in that age subgroup managed in that hospital type.

### 4.3 Independent predictors of operative management

The multivariable logistic regression model initially included the following variables: age, management outside PTC, location/grade of pancreatic injury, ICISS, associated intraabdominal injuries (spleen, hepatobiliary, hollow viscus, other intraabdominal), head injury and transfusion. Age, management outside PTC, ICISS and transfusion were forced into the model. Independent predictors of OM were: high-grade injury (adjusted odds ratio (AOR): 9.18, 95%CI: 2.74-30.69; p<0.0001), hollow viscus injury (AOR: 4.02, 95%CI: 1.23-13.15; p=0.02) and transfusion (AOR: 4.68, 95%CI: 1.27-17.27; p = 0.02) (Fig. 1). Notably, age, ICISS and management outside a PTC were not associated with OM. Further statistical data are available in Appendix C.

### 4.4 Secondary outcomes

Complications occurred in 30.2% (n=42/139) of children (Table 4). One-fifth (n=28/139, 20.1%) of children had pancreatic complications (Table 6). Compared to the NOM group, children who underwent OM were more likely to develop a pseudocyst (n=9/37, 24.3% vs n=4/102, 3.9%, p=0.001). The median hospital LOS was 7 days and was longer in those who underwent OM (19.0 vs 5.0 days, p<0.001). Median ICU LOS was also longer in the OM group (14.0 vs 0 hours, p<0.001). Thirty-one children were readmitted within 90 days, with significantly more in the OM group (n=15/37, 40.5% vs n=16/102, 15.7%, p=0.003). Overall, two children died (1.4%), with one having undergone OM.

**Table 4.** Secondary outcomes by management strategy in children.

|  | Overall | NOM | OM | p-value |
| --- | --- | --- | --- | --- |
| N (%) | 139 (100) | 102 (73.4) | 37 (26.6) |  |
| Any complication | 42 (30.2) | 22 (21.6) | 20 (54.1) | **<0.001** |
| Pancreatic complications | 28 (20.1) | 18 (17.6) | 10 (27.0) | 0.24 |
| Pancreatitis | 18 (12.9) | 16 (15.7) | 2 (5.4) | 0.15 |
| Pseudocyst | 13 (9.4) | 4 (3.9) | 9 (24.3) | **0.001** |
| Other | 1 (0.7) | 0 | 1 (2.7) | 0.27 |
| Other complications | 17 (12.2) | 7 (6.9) | 10 (27.0) | **0.003** |
| AKI | 3 (2.2) | 2 (2.0) | 1 (2.7) | 1.00 |
| ARDS | 0 | 0 | 0 | - |
| Peritonitis | 1 (0.7) | 0 | 1 (2.7) | 0.27 |
| Wound rupture/dehiscence | 2 (1.4) | 0 | 2 (5.4) | 0.07 |
| Wound infection | 6 (4.4) | 1 (1.0) | 5 (13.5) | **0.005** |
| UTI | 5 (3.6) | 3 (2.9) | 2 (5.4) | 0.61 |
| DVT | 1 (0.7) | 1 (1.0) | 0 | 1.00 |
| PE | 0 | 0 | 0 | - |
| Pneumonia | 5 (3.6) | 3 (2.9) | 2 (5.4) | 0.61 |
| Sepsis | 3 (2.2) | 1 (1.0) | 2 (5.4) | 0.17 |
| Stroke | 1 (0.7) | 1 (1.0) | 0 | 1.00 |
| MI | 0 | 0 | 0 | - |
| Paralytic ileus/intestinal obstruction | 6 (4.3) | 4 (3.9) | 2 (5.4) | 0.66 |
| Other | 2 (1.4) | 1 (1.0) | 1 (2.7) | 0.46 |
| LOS days, median (IQR) | 7 (4.0-17.0) | 5.0 (3.0-5.0) | 19.0 (10.5-28.5) | **<0.001** |
| ICU hours, median (IQR) | 0 (0-18.0) | 0 (0-0) | 14.0 (0-84.0) | **<0.001** |
| Ventilator hours, median (IQR) | 0 (0-0) | 0 (0-0) | 0 (0-3.5) | **0.019** |
| 90-day mortality | 2 (1.4) | 1 (1.0) | 1 (2.7) | 0.46 |
| 90-day readmission | 31 (22.3) | 16 (15.7) | 15 (40.5) | **0.003** |

Data are presented as column percentages unless otherwise indicated. Bolded p-values indicate statistical significance. NOM: non-operative management; OM: operative management; AKI: acute kidney injury; ARDS: acute respiratory distress syndrome; UTI: urinary tract infection; DVT: deep vein thrombosis; PE: pulmonary embolism; MI: myocardial infarction; LOS: length of stay; IQR: interquartile range.

## 5. Discussion

### 5.1 Significance of study

To our knowledge, this is the largest Australian study of children with pancreatic injury. Although an uncommon solid organ injury, pancreatic injury is associated with significant morbidity and management remains controversial. There are no established paediatric management guidelines and adult approaches may not be applicable to the paediatric context. In NSW specifically, there are no clear guidelines on where and how children with such injuries should be managed. This provides up-to-date information on patient demographics and injury characteristics, management strategies and outcomes. Most children were managed at a children’s hospital and non-operatively, with lower mortality and comparable morbidity to other studies [5, 7]. Overall, children managed outside PTCs were no more likely to undergo OM, although operation rates were higher at ATCs.

### 5.2 Patient demographics and injury characteristics

Children with BPI were predominantly male, consistent with literature [5, 7, 12, 51–55]. The main mechanisms of injury were pedal cyclist followed by MVA; the predominance of these two mechanisms is consistent with previous studies [5, 27, 52, 55–58], highlighting the need for injury prevention intiatives to be targeted towards these mechanisms. The low incidence of penetrating pancreatic injury in children compared to the US [5] and population-level studies from other countries [51, 59, 60] likely reflects that firearm-related injury in Australia remains uncommon [61–63].

Almost two-thirds of children with BPI were managed at a PTC. In comparison, a 2017 study on paediatric blunt splenic injury in NSW found that only one-third of cases were admitted or transferred to a PTC [37]. This is echoed by international studies on paediatric trauma more generally that report 63-89% of children being managed outside PTCs [38, 43, 45, 47, 64–66]. The reasons that children with pancreatic injury may be more likely to be managed at a PTC are unknown. However, there was a much higher rate of associated injuries compared to the NSW study on blunt splenic injury, which may possibly explain this difference. It is also possible that there is relatively less comfort with managing children with pancreatic trauma compared to other trauma in non-paediatric settings.

### 5.3 Management of children with blunt pancreatic injury in New South Wales

Three-quarters of children with BPI were managed non-operatively, with no association between management at PTCs versus non-PTCs overall and the management strategy. While two-thirds of cases at ATCs (n=8/12, 66.6%) were managed operatively, this needs to be interpreted with caution: it represents only eight operations over 18 years.These small numbers meant that individual centre types could not be included in the regression model to allow adjustment for possible confounders. However, it does echo findings from a previous study on paediatric splenic injury in NSW, where ATCs had the highest rate of OM [37]. It is possible that adherence to adult pancreatic injury management guidelines, which recommend OM where ductal injury is suspected, explains this disparity. Further research to verify and explore possible explanations is required.

Notably, the small number of children undergoing a pancreas-specific operation (7.9%) is less than a third of that reported in other studies. Englum et al. conducted a similar but larger (n=674) study using the United States National Trauma Data Bank and found that 23.7% of children with BPI underwent a pancreas-specific operation [5]. Other single and multicentre studies from the US have reported laparotomy rates of 24.9-54.5% [53, 56] and pancreatic operation rates of 34.1-40.5% [27, 53, 54]. Additionally, only 2.2% of the children in our study had a splenectomy, compared to 14.2% reported by Englum et al. [5]. This is positive as splenectomy should be avoided if possible due to the life-long increased risk of infections and the associated mortality, particularly in children [67]. Reasons for these different findings are unclear, however, with no higher morbidity in our study, avoiding an operation on the pancreas is supported. Further exploration of the drivers of paediatric pancreatic injury management decisions in different contexts is required. Given the higher rates of OM for other solid organ injuries away from PTCs [37, 46, 68-73) the higher rates of OM in ATCs for both pancreatic and splenic injury in NSW [37], and the improved outcomes reported at PTCs for paediatric trauma more generally [38–43], there may be the potential to develop clinical practice guidelines to support the care of children with pancreatic injury; specifically, this may include defining the place of NOM and creating more nuanced transfer protocols.

#### 5.3.1 Other independent predictors of operative management

Other factors correlated with OM on multivariable analysis included high-grade injury, transfusion as a proxy for haemodynamic instability and hollow viscus injury. Whilst high-grade BPI was associated with OM, only 40% of children with high-grade injury underwent an operation. Englum et al. also reported that pancreatic injury grades IV and V predicted a pancreas-specific operation, although interestingly this association did not hold for grade III injury (distal high-grade injury) [5]. These findings reflect the ongoing debate about the management of high-grade BPI.

In guidelines for the management of other solid organ injuries, haemodynamic instability has replaced injury grade as the indication for OM [15–17, 74–78]. In contrast, damage to the pancreatic duct is the primary indication for operation in pancreatic injury as it is the main determinant of morbidity [19, 20]. The association between transfusion and OM might be explained by the fact that over 70% of children undergoing OM had other intraabdominal injuries.

The association between hollow viscus injury and OM is not surprising as they almost always require an operation due to peritoneal contamination [79] and the high morbidity and mortality associated with a delay in management [80, 81]. Indeed, the shift to NOM for solid organ injuries means that a hollow viscus injury is now one of the commonest reasons for a laparotomy after blunt abdominal trauma [82]. Of note, overall injury severity was not associated with OM. Englum et al. [5] as well as studies of other solid organ injuries [48, 65, 83] have shown that higher injury severity or the presence of multiple injuries is associated with OM, whilst others have found no association [37].

### 5.4 Secondary outcomes

The mortality of 1.4% in this study was low compared to other studies [5, 7], supporting the non-operative approach. Morbidity was 30% overall, with 20% having a complication specific to the pancreas. Englum et al. reported a similar morbidity rate of 26.5%, although they did not account for pancreas-specific complications [5]. These numbers reflect the high morbidity associated with pancreatic injury although it remains unclear how this is affected by the management approach. Wound infection aside, pseudocyst formation was the only complication observed to be significantly different between OM and NOM groups; surprisingly, this was much lower in those managed non-operatively. This is difficult to interpret given the small numbers and the fact that OM did not necessarily equate to having a procedure on the pancreas. Others have suggested that pseudocyst formation is more common in those undergoing NOM, with an incidence of 8-45% [7, 23, 56, 84, 85] compared to less than 2% in OM [7, 85]. No cases in our study developed a pancreatic fistula – which others have found to be more common after OM [22].

The risk of pseudocyst following NOM of high-grade pancreatic injury has traditionally been considered an argument for OM [25]. However, endoscopic drainage via cyst-gastrostomy appears to be effective in managing them and may lead to overall lower morbidity [23, 24]. Furthermore, early endoscopic stenting for minor ductal disruptions in selected haemodynamically stable patients may reduce the risk of pseudocyst and remove the need for OM altogether [23, 24, 53, 54]. The technical challenges of this procedure in a paediatric population, with the subsequent risk of ductal stricture remains a concern [86]: most studies on this topic are case reports, with the few case series being retrospective, heterogeneous and having small sample sizes [25].

### 5.5 Limitations of this study

Being retrospective and using administrative data not conceived for research purposes, this study was limited by information bias. There was no control group or ability to adjust for unmeasured confounders including physiological parameters, clinical progress and intraoperative or radiological findings. The drivers of surgeon decision-making were also unknown. The study leverages 18 years of data, during which management practices may have changed. Defining pancreatic grade according to the Organ Injury Scale was difficult as the ICD-10-AM does not specify pancreatic injury grade. Nevertheless, the ICD-10-AM definition of high-grade injury used was predictive of OM in the multivariable analysis, suggesting that it reflects pancreatic injury severity. Similarly, whilst the Abbreviated Injury Scale (AIS) has been regarded as the standard for injury coding, mapping of AIS to ICD-10-AM diagnoses was unavailable. ICISS was used to indicate injury severity, although this also has several limitations [87].

Regarding our statistical analysis, small numbers in certain variable groups such as age group and hospital type limited interpretation of results. The small sample size also limited the number of variables in the multivariable analysis. Notably, we did not adjust for mechanism of injury. We were also unable to specifically examine the management of children at ATCs, where operation rates were higher. It is possible that with larger numbers and the ability to adjust for management at ATCs independently, this may be found to be an independent predictor of OM in children.

Finally, the definition of OM as any abdominal operation was inclusive of non-pancreatic operations, although it did correlate with pancreatic injury severity. We were also unable to examine whether the presence of a BPI influenced surgical decision-making in those patients who did not undergo a pancreas-specific operation.

## 6. Conclusion

This is the largest Australian study of paediatric BPI. Operation rates were lower than reported internationally, with comparable morbidity and lower mortality. These results support a non-operative approach to blunt pancreatic injury in children in most cases. While the rate of OM at ATCs was higher than at other centres, treatment outside a PTC was not associated with the likelihood of operation after adjusting for confounders. The large number of children with BPI managed at a PTC compared to other solid organ injuries may indicate the perceived complexity of such injuries and the associated management decisions. However, there is a paucity of knowledge regarding the drivers of such management decisions, including operation and transfer to a PTC. Further research is required to provide nuanced guidance on the management of pancreatic injury, especially high-grade injury, and regarding the need for transfer.

## Declarations of interest

none

## Funding

none

## Data Availability

All data produced in the present study are available upon reasonable request to the authors

## Notes

### Competing Interest Statement

The authors have declared no competing interest.

### Funding Statement

This study did not receive any funding

### Author Declarations

NSW Population and Health Services Research Ethics Committee gave ethical approval for this work.

